# Sensitive detection of tumor mutations from blood and its application to immunotherapy prognosis

**DOI:** 10.1101/2019.12.31.19016253

**Authors:** Shuo Li, Zorawar Noor, Weihua Zeng, Xiaohui Ni, Zuyang Yuan, Frank Alber, Wenyuan Li, Edward B. Garon, Xianghong Jasmine Zhou

**Affiliations:** Department of Pathology and Laboratory Medicine, David Geffen School of Medicine, University of California at Los Angeles, Los Angeles, CA 90095, USA; Bioinformatics Interdepartmental Graduate Program, University of California at Los Angeles, Los Angeles, CA 90095, USA; Department of Medicine, Division of Hematology/Oncology, David Geffen School of Medicine at UCLA, Los Angeles, CA 90095, USA; EarlyDiagnostics Inc., Los Angeles, CA 90095, USA; Department of Microbiology, Immunology and Molecular Genetics, University of California at Los Angeles, Los Angeles, CA 90095, USA

## Abstract

Liquid biopsy using cell-free DNA (cfDNA) is attractive for a wide range of clinical applications, including cancer detection, locating, and monitoring. However, developing these applications requires precise and sensitive calling of somatic single nucleotide variations (SNVs) from cfDNA sequencing data. To date, no SNV caller addresses all the special challenges of cfDNA to provide reliable results. Here we present *cfSNV*, a revolutionary somatic SNV caller with five innovative techniques to overcome and exploit the unique properties of cfDNA. *cfSNV* provides hierarchical mutation profiling, thanks to cfDNA’s complete coverage of the clonal landscape, and multi-layer error suppression. In both simulated datasets and real patient data, we demonstrate that *cfSNV* is superior to existing tools, especially for low-frequency somatic SNVs. We also show how the five novel techniques contribute to its performance. Further, we demonstrate a clinical application using *cfSNV* to select non-small-cell lung cancer patients for immunotherapy treatment.

## Introduction

Cell-free DNA (cfDNA) in blood has received enormous attention thanks to its clinical utility as a surrogate for tumor biopsy, especially in cases where the latter is unavailable or insufficient [1]. A tissue biopsy is invasive by nature, and is only extracted from a single site. In contrast, cfDNA in blood can be obtained noninvasively, and provides a comprehensive landscape of the heterogeneous genetic alterations in tumors. Hence, a wide range of cfDNA-based applications have been developed to detect cancer [2-5], locate tumors in the body [4,6], select the best therapy [7,8], and monitor treatment [6,9,10]. All these applications depend upon an indispensable, yet underdeveloped task: precise and sensitive calling of somatic single nucleotide variations (SNV) from cfDNA sequencing data. This task is challenging to conventional SNV callers because **somatic mutations in cfDNA generally have low prevalence. This property follows from the major hallmarks of cfDNA:** (1) cfDNA is a mixture of DNA fragments from both normal and tumor cells, and in most cancer patients the fraction of tumor-derived cfDNA is extremely low (<1% for most early-stage cancer patients [11] and <10% even for some metastatic patients [12]). (2) cfDNA comes from the entire volume of a tumor, and from every tumor present in a patient, so it provides complete information on clonal and subclonal mutations.

Existing methods are not equipped to handle this complicated scenario. Specifically, they are lacking in three aspects: (1) They do not account for the low fraction of tumor-derived cfDNA or variability due to the tumor clonal hierarchy. A few SNV callers (e.g., *MuTect* [13]) try to handle the issue of tumor impurity, but even these cannot robustly and sensitively detect mutations with variant allele frequency (VAF) <5% [13]. (2) Their post-filtration steps require the reliable estimation of site-level statistics (e.g. strand bias and averaged base quality). Robust estimates are already challenging to obtain for low-frequency cfDNA mutations, due to insufficient supporting variant reads, and become even more difficult for whole-exome sequencing (WES). WES is increasingly popular, but does not permit deep sequencing. (3) They do not exploit two key features of cfDNA, namely short fragment size (∼166 bp on average) and non-random fragmentation [14, 15].

Therefore, we have developed a revolutionary cfDNA SNV caller named *cfSNV*. This is the *first* algorithm to address all the cfDNA-specific challenges and opportunities mentioned above, taking advantage of modern statistical models and machine learning approaches. *cfSNV* achieves high precision and sensitivity in both low-purity and highly heterogeneous cfDNA samples, even with medium-coverage sequencing data such as WES. In contrast, existing cfDNA SNV callers address these challenges only partially and rely on specially designed experimental techniques (e.g. ultra-deep and/or barcode-based sequencing [16, 17]). Therefore, our method has a broad spectrum of applications in cancer detection, cancer monitoring, and treatment selection. As an example application, in this study we demonstrate that *cfSNV* can facilitate the noninvasive selection of non-small-cell lung cancer patients for immunotherapy treatment.

## Results

### *cfSNV*: A new computational framework for calling SNVs from cfDNA

We developed the *cfSNV* framework (Fig. 1c) by introducing five new techniques (Fig.1b) into the standard SNV calling workflow (Fig. 1a). Each of the five techniques either overcomes a specific challenge of cfDNA or takes advantage of a specific feature of cfDNA. The challenges and features are:

**Fig. 1.**
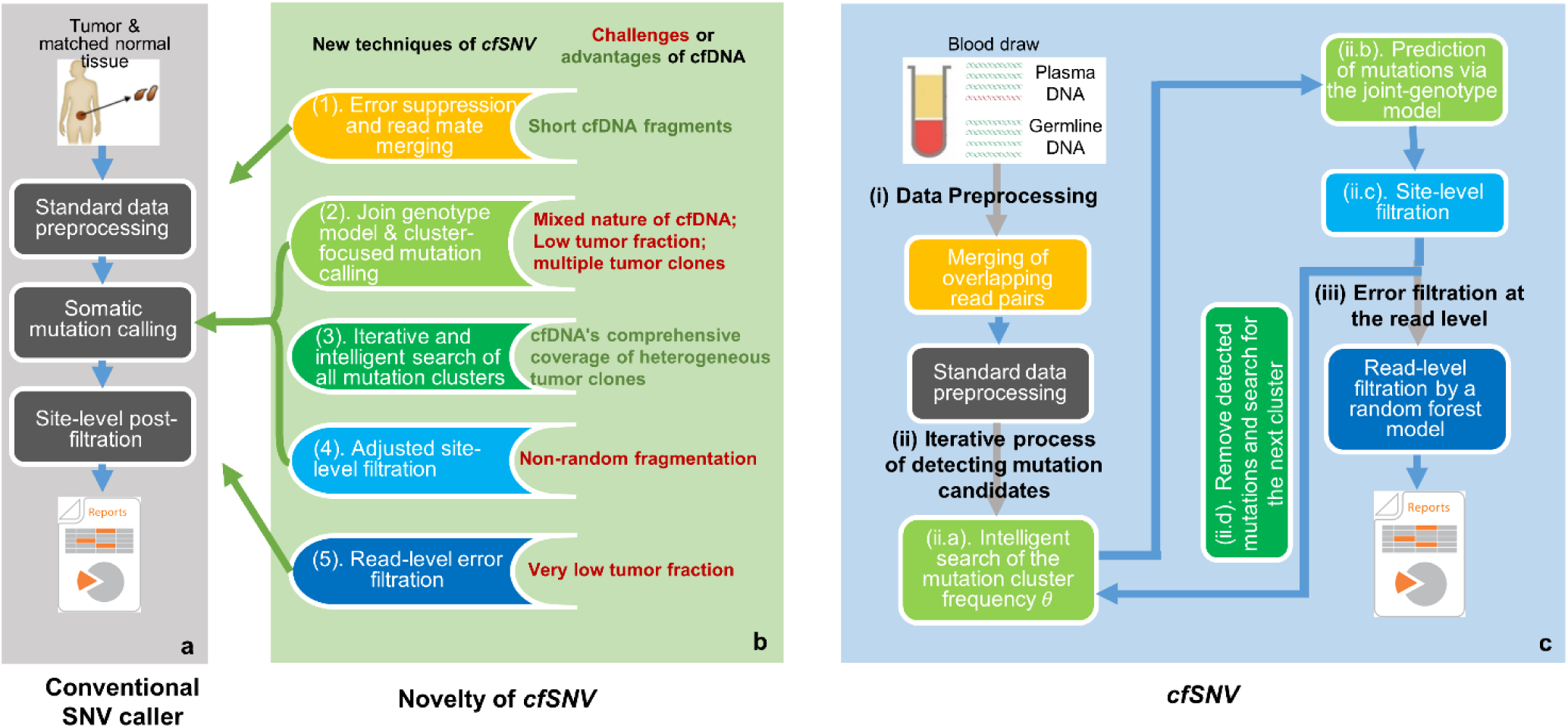
*cfSNV* framework and its novel techniques. **a**, The workflow of conventional SNV callers takes the genomic data of a tumor and its matched normal tissue as inputs. **b**, Five new techniques introduced to *cfSNV* that modify the standard workflow. **c**, Full workflow of *cfSNV. cfSNV* takes plasma DNA and germline DNA sequencing data as inputs. It first merges overlapping read pairs in cfDNA sequencing data. Next, we apply standard data preprocessing tools. An iterative procedure then detects mutation clusters and estimates their frequencies θ based on multiple, automatically selected hotspots. Each iteration determines joint genotypes across sequencing regions to predict somatic SNV candidates, and masks the mutation candidates before proceeding. After all clusters and mutation candidates have been detected, a random forest classifier identifies raw read pairs with sequencing errors. Finally, somatic SNVs are reported only if enough variant supporting read pairs passed the random forest screening. The background color of steps in **c** corresponds to the feature listed in **b**.

#### (1) Short fragments

the fragment length distribution of cfDNA peaks at 166bp. Therefore, paired-end sequencing (usually 150 bp for a read) usually results in a large fraction of overlapping read mates, which can be used to suppress sequencing errors (Fig. 1b(1) and Fig. 1c(i)). This error-correction step is performed before the standard data preprocessing.

#### (2) Mixed nature

the cfDNA found in blood from cancer patients generally consists of a small amount of tumor-derived cfDNA among an overwhelming majority of cfDNA from normal cells. By incorporating the germline data of white blood cells (WBCs) from the same subject, we can fit a joint-genotype model that precisely describes this mixture. Specifically, we model the triplet (*g*_*T*_, *g*_*N*_, *g*_*W*_) of genotypes from tumor-derived cfDNA, normal-derived cfDNA, and matched WBC DNA. This is done by first aggregating reads from mutation hotspots in order to robustly estimate the tumor-derived cfDNA fraction, then applying our joint-genotype model to probabilistically deconvolute tumor- and normal-derived reads in a specific locus.

#### (3) Heterogeneous clonal compositions

unlike tissue biopsies, a blood sample includes DNA fragments from all tumor sites, so it covers the full range of clonal and subclonal mutations [11, 18]. However, admitting a heterogeneous cfDNA clonal composition poses a great challenge to existing methods. A statistical model capable of fitting the data from clonal mutations, inevitably sacrifices accuracy for subclonal mutations using the same parameters. We can take advantage of the fact that the mutations associated with a given clone have similar prevalence in cfDNA. The mutations are therefore naturally clustered according to the clonal hierarchy [11, 18]. This fact permits us to develop a “divide-and-conquer” algorithm (Fig. 1c(ii)) that first automatically groups the heterogeneous mutations into clusters, then fits the model parameters to each cluster of mutations separately. In other words, this algorithm intelligently and iteratively searches for the best parameters of the cfDNA joint-genotype statistical model (Fig. 1c(ii.a)) to detect and model the cluster of mutations with the highest prevalence in the cfDNA sample (Fig. 1c(ii.b)), then removing its loci and data. The process repeats, detecting the next most prevalent mutation cluster at each iteration (Fig. 1c(ii.d)), until no more mutations are detected with confidence. Therefore, we can profile the cfDNA mutation hierarchy in terms of mutation frequencies.

#### (4) Non-random fragmentation

cfDNA fragments have preferred start and end positions [14], so true mutations could cluster at certain positions on the supporting reads. Conventional tools which assume randomly fragmented genomic DNA tend to classify mutation candidates with clustered positions as misalignment artifacts, eliminating them. We remove this artifact filter to keep true cfDNA mutations, while building a new filter to jointly analyze the positions of multiple nearby mutation candidates and precisely remove cfDNA misalignment artifacts (Fig. 1b(4) and Fig. 1c(ii.c)). The new filter successfully rescued 1∼16 mutations (median 6.5) per subject that would have been discarded by conventional methods.

#### (5) Confusion between sequence errors and low-frequency mutations

When the tumor-derived cfDNA fraction is low, sequencing errors impair the detection sensitivity. We get around the problem of low signal-to-noise ratio for individual alleles by developing a machine learning approach to accurately distinguish true variants from sequencing errors for individual reads. The algorithm exploits a variety of contextual information from the region surrounding the target allele (Fig. 1b(5) and Fig. 1c(iii)) to provide an accurate prediction. The detailed workflow is illustrated in Fig. 5 and described in Methods.

### Validation of *cfSNV* on simulation data

To evaluate the performance of *cfSNV* in calling low-frequency somatic mutations, we tested the method on simulated data. To generate the dataset, a set of predefined somatic SNVs were added to the simulation data, the mixture of the germline sequencing data from 11 subjects (around 2000x, see Methods). We used seven variant allele frequencies (VAF) ranging from 0.1% to 5% for the SNVs, in order to simulate tumor heterogeneity among subjects (Methods). Mutations called at positions other than the ground-truth SNVs were regarded as false positives. The results of this test show that *cfSNV* far outperforms two competing methods (*MuTect* and *Strelka2*, both established SNV callers) for all ground-truth mutations (Table 1a). Specifically, *cfSNV* achieves much higher sensitivity (70%) than *MuTect* (23%), *Strelka2* (19%), and *Strelka2* with disabled filters (23%), while maintaining very high precision (99% vs. 92%, 100%, and 22% respectively). When looking at low-frequency mutations specifically, the contrast between *cfSNV* and other methods is even stronger (Table 1b). In this sub-population, *cfSNV* detected 45%∼82% of mutations with VAFs of 0.1%∼1% respectively, whereas most competing methods detected zero mutations. Only *MuTect* achieves a moderate sensitivity: 23% for those mutations with a VAF of 1%.

**Table 1.** Validation of *cfSNV* on simulation data.

| Sub-table a. Performance<sup>§</sup> of *cfSNV*, evaluated based on all ground-truth mutations (561) |  |  |  |  |  |
| --- | --- | --- | --- | --- | --- |
|  | *cfSNV* | *MuTect* | *Strelka2* * | *Strelka2* (filters disabled) * |  |
| # predicted positives | 399 | 139 | 107 | 579 |  |
| # true positives | 395 | 129 | 107 | 127 |  |
| # false positives | 4 | 10 | 0 | 452 |  |
| Sensitivity = $\frac{\text{\#true positives}}{\text{\#ground-truth mutations}}$ | 70% | 23% | 19% | 23% | |
| Precision = $\frac{\text{\#true positives}}{\text{\#predicted positives}}$ | 99% | 92% | 100% | 22% | |
| Sub-table b. Sensitivity of *cfSNV* for mutations with different variant allele frequencies (VAF) |  |  |  |  |  |
| VAF for simulated mutations | # Ground-truth mutations | Sensitivity = $\frac{\text{\#true positives}}{\text{\#ground-truth mutations}}$ (# true positives) | | | |
|  |  | *cfSNV* | *MuTect* | *Strelka2* * | *Strelka2* (filters disabled) * |
| 0.1% | 120 | 45% (54) | 0% (0) | 0% (0) | 0% (0) |
| 0.3% | 121 | 60% (73) | 0% (0) | 0% (0) | 0% (0) |
| 0.5% | 68 | 74% (50) | 3% (2) | 0% (0) | 0% (0) |
| 0.8% | 67 | 75% (50) | 13% (9) | 0% (0) | 0% (0) |
| 1% | 61 | 82% (50) | 23% (14) | 0% (0) | 3% (2) |
| 3% | 64 | 94% (60) | 80% (51) | 84% (54) | 98% (63) |
| 5% | 60 | 97% (58) | 88% (53) | 88% (53) | 100% (60) |
| Total | 561 | 70% (395) | 23% (129) | 19% (107) | 23% (127) |
| <sup>*</sup> *Strelka2* had no false positives because its background scoring model uses a high cutoff and hence sacrifices sensitivity (it has the lowest, 19%), which is not the preferred strategy. To make a fair comparison with the other methods, we disabled the filters of *Strelka2* and achieved a slightly improved sensitivity (23%) but the precision dropped to 22%. *Strelka2* without filters had over 450 false positives — 100 times more than *cfSNV* — yet still had a sensitivity 3 times lower than *cfSNV*. |  |  |  |  |  |
| <sup>§</sup> There are millions of non-mutation loci that are ground truth negatives in this simulation data. Due to the extreme imbalance between positives and negatives, the common performance measure “specificity” (or false positive rate: $\frac{\text{\#false positives}}{\text{\#non-mutation positions}}$) is not suitable for comparing the methods. This is because it uses a fixed huge number in the denominator and a small number in the numerator. | | | | | |

### Validation of *cfSNV* on patient data

Next, we tested the ability of *cfSNV* to call somatic mutations on real patient data. We collected WES data of samples obtained from six metastatic prostate cancer (CRPC) and twelve metastatic breast cancer (MBC) patients [19] (Methods). For each patient, we collected a metastatic tumor biopsy sample, a WBC sample, and two plasma cfDNA samples. The cfDNA samples were drawn at two different time points after the patients were diagnosed as metastatic, with time gaps in the range 14∼138 days. We compare the different cfDNA SNV callers in terms of the fraction of mutations detected in one cfDNA sample that are also detected as variant supporting reads in either the matched tumor tissue or the other cfDNA sample (this fraction is defined as the “confirmation” rate, see Methods). The confirmed mutations are regarded as true positives. As all tumor clones in these metastatic cancer patients can hardly be profiled from a single tumor biopsy sample, we regarded mutations found in both plasma samples and not the tumor biopsy also as true mutations from unprofiled clones.

First, we tested the confirmation rate of *cfSNV* across different samples. We applied *cfSNV* to the 18 initial cfDNA samples to obtain a baseline mutation set for calculating the confirmation rate. We validated the truncal and branch mutations detected. A mutation was defined as “truncal” if the VAF was above 60% of the highest VAFs in the sample and was defined as “branch” otherwise (Methods). Averaged across subjects, 97.7% and 76.7% of truncal mutations are confirmed in the later cfDNA sample and the tumor biopsy respectively. 93.2% and 62.1% of branch mutations are confirmed in the later cfDNA sample and the tumor biopsy respectively (Fig. 2a). The confirmation rates are similar if we instead use mutations detected in the 18 later cfDNA samples as a baseline (Fig. 2a, 96.5% and 78.6% for truncal mutations, 93.0% and 59.9% for branch mutations in the earlier cfDNA sample and the tumor biopsy respectively). As time goes, the mutation landscape appearing in cfDNA changes, especially for branch mutations. Reflecting this fact, we observe that the larger the time gap between the two blood draws, the lower the confirmation rate of branch mutations between the two cfDNA samples (Pearson’s correlation between the time gap and the confirmation rate = –0.51, p = 0.034). The existence of this trend can lead us to underestimate the performance of *cfSNV* by these kinds of confirmation tests. Across cancer types, metastatic breast cancer has a higher tissue-based confirmation rate than metastatic prostate cancer (truncal 83.4% vs. 66.2%, branch 64.5% vs. 54.0%), but the two cancer types have similar cfDNA-based confirmation rates (truncal 96.2% vs. 98.9%, branch 93.1% vs. 93.1%). This result may be attributed to the fact that the circulation level of breast cancer is higher than that of prostate cancer [20].

**Fig. 2.**
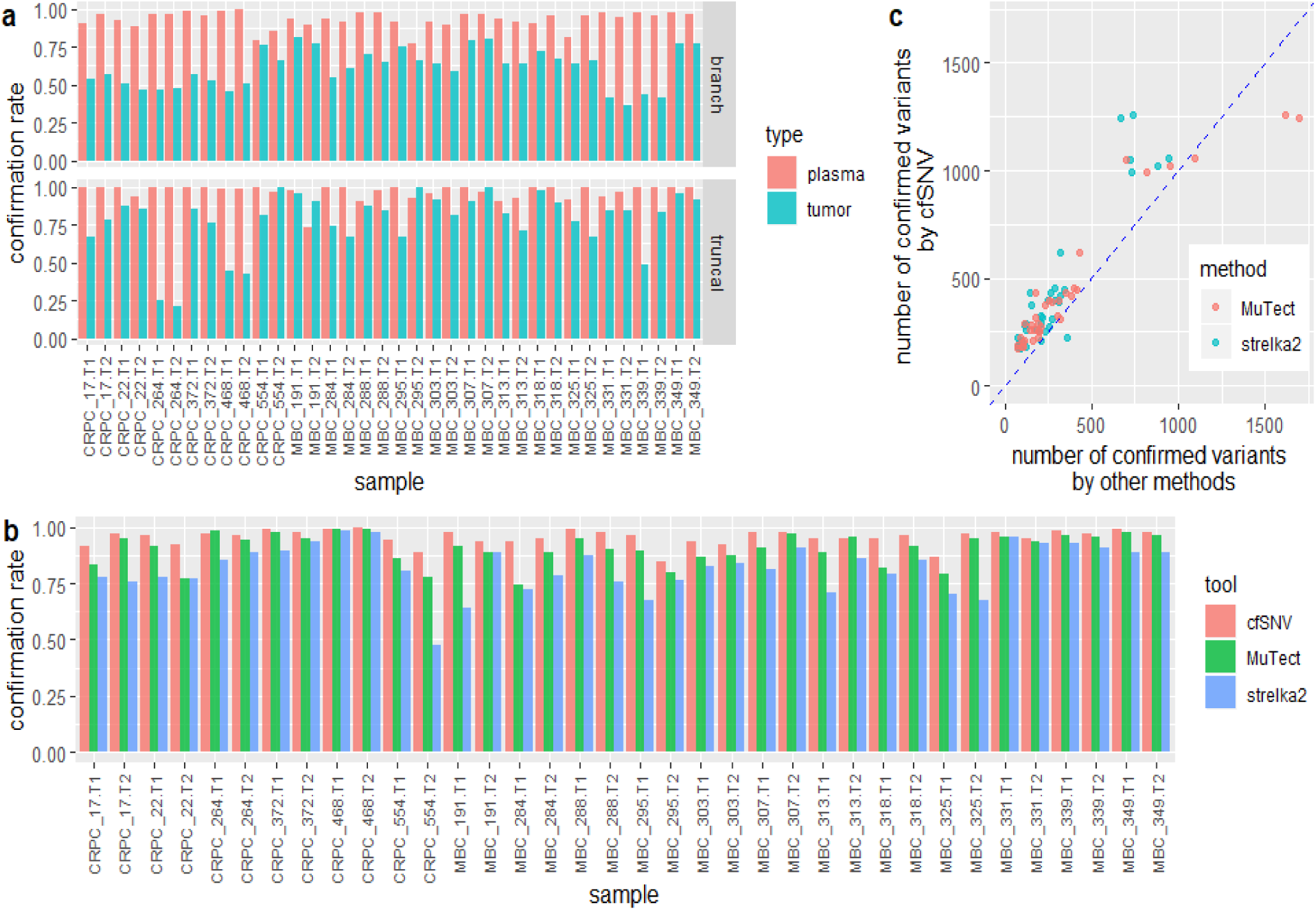
Somatic SNV calling on cfDNA sequencing samples from cancer patients. **a**, Fraction of confirmed truncal mutations and branch mutations detected by *cfSNV*. Mutations found in cfDNA sequencing data were validated by variant supporting read counts, either in cfDNA sequencing data from the other plasma sample or in genomic DNA sequencing data from a tumor biopsy sample collected from the same patient. The clonality of mutations was determined by their relative VAFs. **b**, Fractions of confirmed somatic SNVs found by *cfSNV, MuTect* and *Strelka2*. The fraction of confirmed somatic SNVs is the number of confirmed mutations divided by the total number of reported mutations. In the sample name, “T1” and “T2” indicate the first time point and the second time point of blood plasma samples respectively. **c**, Total number of confirmed variants found by *cfSNV, MuTect* and *Strelka2*.

Second, we compare *cfSNV* with the confirmation rates obtained by competing methods (*MuTect* and *Strelka2*) on the same samples. *cfSNV* outperformed both methods, achieving the highest confirmation rate (a mutation is confirmed if it is found in either the tumor sample or the other cfDNA sample) in 33 out of 36 samples (Fig. 2b). For the remaining 3 samples, *cfSNV*’s confirmation rate was only marginally lower than the highest confirmation rate (by 0.2%, 0.9%, and 1.2%). This result shows that the mutations detected by *cfSNV* are more likely to be true positives, which translates into higher precision. Moreover, *cfSNV* detected more confirmed mutations overall (Fig. 2c), indicating higher sensitivity. Even if the confirmation rate in the tumor sample were the only criterion, *cfSNV* performs still better than or comparably to the competing methods in terms of sensitivity and precision (Supplementary Fig. 1). However, it is also important to notice that all three methods have consistently higher confirmation rates in the second plasma sample than the matched tumor tissue sample, implying that plasma cfDNA offers a more comprehensive coverage of tumor mutations than a single tumor biopsy for advanced cancer patients. Whenever multifocal sampling of tumors from a metastatic cancer patient is infeasible, cfDNA is a viable alternative to obtain comprehensive mutation profiles.

### Application to predict the outcome of anti-PD-1 treatment: a new bTMB measure

Recent studies [8, 21] have shown that bTMB is an attractive alternative to TMB due to three advantages: (1) noninvasiveness, (2) more comprehensive mutation coverage (by cfDNA) than a single-site tumor biopsy, and (3) the VAFs of mutations reflects their clonality. It has also been shown that high truncal neoantigen load and low intra-tumor heterogeneity, more significantly associate with longer progression-free survival (PFS) [22, 23] than total neoantigen load alone. To fully exploit advantages (2) and (3), WES profiling of cfDNA is needed. Due to the lack of efficient tools to accurately call SNV from cfDNA using medium-coverage WES data (e.g. 200x), current bTMB methods [8, 21] all use small gene panels (<600) in order to perform deep sequencing (e.g. >5000x). Small panels can only sparsely sample the total mutation landscape, so the resulting estimates of TMB or bTMB are influenced by population and sampling variation [24]. In contrast, *cfSNV* fully profiles the mutation landscape as well as allowing us to benefit from all the other advantages offered by cfDNA. Specifically, we have developed a new immunotherapy prognosis metric, truncal-bTMB, which uses only truncal mutations called by *cfSNV* from the WES profiling of cfDNA samples (Methods). We applied this new measure to predict the outcomes of anti-PD-1 treatment, and achieved superior performance compared with TMB and bTMB.

To evaluate the predictive power of the three measures (TMB, bTMB, and truncal-bTMB), we studied a cohort of 30 non-small-cell lung cancer patients who received anti-PD-1 treatment (pembrolizumab). Blood samples were drawn from them before treatment. Fourteen of the patients also had tumor biopsies (Methods). First, we compared all three measures for the 14 patients with both tumor biopsies and cfDNA samples. Next, we compared bTMB with truncal-bTMB performed on the cfDNA samples of all 30 patients. All samples were sequenced with WES. We split the 14 patients with tumor biopsies into two groups around the population median [25] of each measure, which we call the high-burden and low-burden groups. We then evaluated the performance of the measures based on how well they separate the Kaplan-Meier survival curves of the high-burden and low-burden groups (PFS). Our truncal-bTMB method had the most significant one-sided log-rank test p-value, 0.028 (truncal-bTMB) vs. 0.280 (TMB) and 0.067 (bTMB), implying that it has the highest power for predicting patients with longer PFS. Those patients are the most likely benefit from immunotherapy (Fig. 4a-c). We conducted a similar experiment with all 30 patients, this time comparing only bTMB and truncal-bTMB; again, truncal-bTMB performs better (Fig. 4d-e, p-value 0.012 for truncal-bTMB vs. 0.069 for bTMB). Note that truncal-bTMB actually showed higher significant difference between survival curves in both experiments than bTMB, indicating that combining mutation clonality and intra-tumor heterogeneity improves predictive power. Therefore, our proposed new measure exploits the unique advantages of cfDNA to providing a superior anti-PD-1 prognosis indicator.

### Experimental analysis of five new techniques

Here, we quantitatively assess how each of the five new techniques contributes to the performance of *cfSNV*.

#### (i) Suppression of sequencing errors using overlaps of read mates

The pair-end sequencing of cfDNA results in significant overlaps in the read mates. For example, in 95% of 59 cfDNA samples collected from *Adalsteinsson et al*. [19], >50% of read mates overlap (Supplementary Fig. 2). Our result shows that using overlapping read pairs, combined with a machine learning approach (see point (v) below and Fig. 1b(5)), can greatly facilitate the detection of true mutations while rejecting sequencing errors. Specifically, if we compare a model using the overlapping read information to one that does not, the AUC performance averaged across 12 out-of-sample test datasets (cfDNA samples from *Adalsteinsson et al*. [19]) shows significant improvement (Wilcoxon rank sum test p-value = 0.02, Supplementary Fig. 2).

#### (ii) Enhanced mutation detection by a joint-genotype model parameterized by the frequency *θ*, which serves as a proxy for the VAFs of mutations in the most prevalent mutation cluster

This facet of our technique can be assessed in two ways *(1) Does θ actually reflect the VAFs of the mutations in the most prevalent mutation cluster*? To answer this question, we designed three experiments using simulated data with synthetic mutations, simulation data obtained by mixing real sample data with a known dilution ratio, and real cfDNA data. In the first experiment, we generated sequencing data with three groups of synthetic mutations: one mutation cluster with a VAF of 20%, one cluster with a VAF of 8%, and one with a VAF of 2% (Methods). Our method not only automatically identifies the most prevalent cluster and estimates its VAF, but also finds the other two clusters in subsequent iterations (Supplementary Fig. 3). In the second experiment, we subsampled and mixed sequencing reads from WBC and cfDNA samples, both taken from the same cancer patient (Methods). The tumor fraction, which is estimated by the frequency *θ* of the most prevalent mutation cluster in these mixed samples, correlates very strongly (Pearson’s correlation = 0.99) with the ground-truth mixing dilution (Fig. 4a) across the study population. In the third experiment, we used data from two independent sequencing experiments (WES and WGS) on the same cfDNA sample from cancer patients. Specifically, we compare the tumor fraction estimated by *cfSNV* on WES to that estimated by *ichorCNA* on WGS. This result, shown in Supplementary Fig. 3, also confirms that our method accurately estimates the prevalence of the major mutation clusters. *(2) Does accurately estimating the mutation cluster frequency θ enhance mutation detection?* To test this facet of *cfSNV*, we generated simulated sequencing data for a list of predefined *θ* values, from 0% to 100%, and observed the optimal *θ* that fits the joint-genotype model. Our performance metric is the model-to-data fitness ratio, defined as the ratio between the likelihoods of correct and incorrect joint genotypes (Methods). A higher ratio means that the model is a better fit, so the mutation is more likely to be identified. Our result shows that any given mutation is best fit by the model when *θ* takes on a value close to the mutation’s prevalence (Supplementary Fig. 3). In addition, when comparing the fitness of the model with and without *θ* (i.e., comparing the two likelihood ratios), we find that the smaller a mutation’s VAF, the larger the difference (e.g., the model-to-data fitness ratio is 40 times higher with *θ* present, for VAF<5%). This relationship indicates that an accurate *θ* estimate significantly enhances the detection power for low-frequency mutations (Fig. 4b). Furthermore, we used cfDNA samples whose prevalent mutation clusters have low prevalence (<20%) to further confirm this conclusion (Supplementary Fig. 3). More mutations were detected when the assigned *θ* approached the true value of the mutation cluster frequency.

**Fig. 3.**
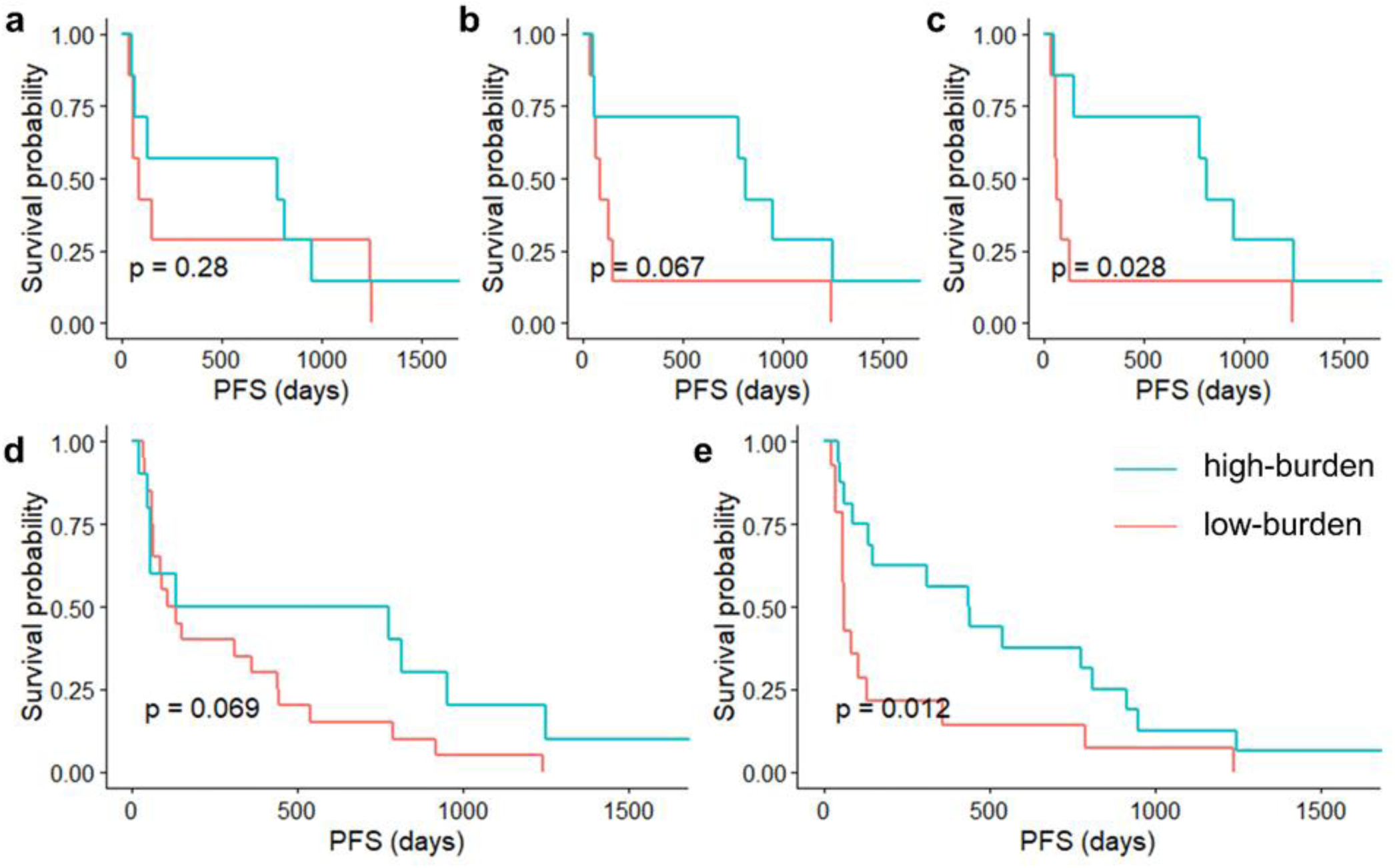
Kaplan-Meier curves for progression-free survival (PFS) on advanced non-small cell lung cancer patients. **a-c**, PFS for 14 patients with both tumor biopsy and pre-treatment cfDNA sequencing data. The high-burden and low-burden groups in each plot are defined by the median value of the measure: TMB (**a**, HR=0.721, 95% CI [0.239, 2.173]), bTMB (**b**, HR=0.411, 95% CI [0.124, 1.355]), or truncal-bTMB (**c**, HR=0.326, 95% CI [0.098, 1.079]). **d-e**, PFS curves for all 30 patients with pre-treatment cfDNA sequencing data. **d**, The high-burden and low-burden groups are determined by the bTMB cutoff value from the 14 patients (HR=0.530, 95% CI [0.226, 1.244]). **e**, The high-burden and low-burden groups are determined by the truncal-bTMB cutoff value from the 14 patients (HR=0.425, 95% CI [0.199, 0.907]).

**Fig. 4.**
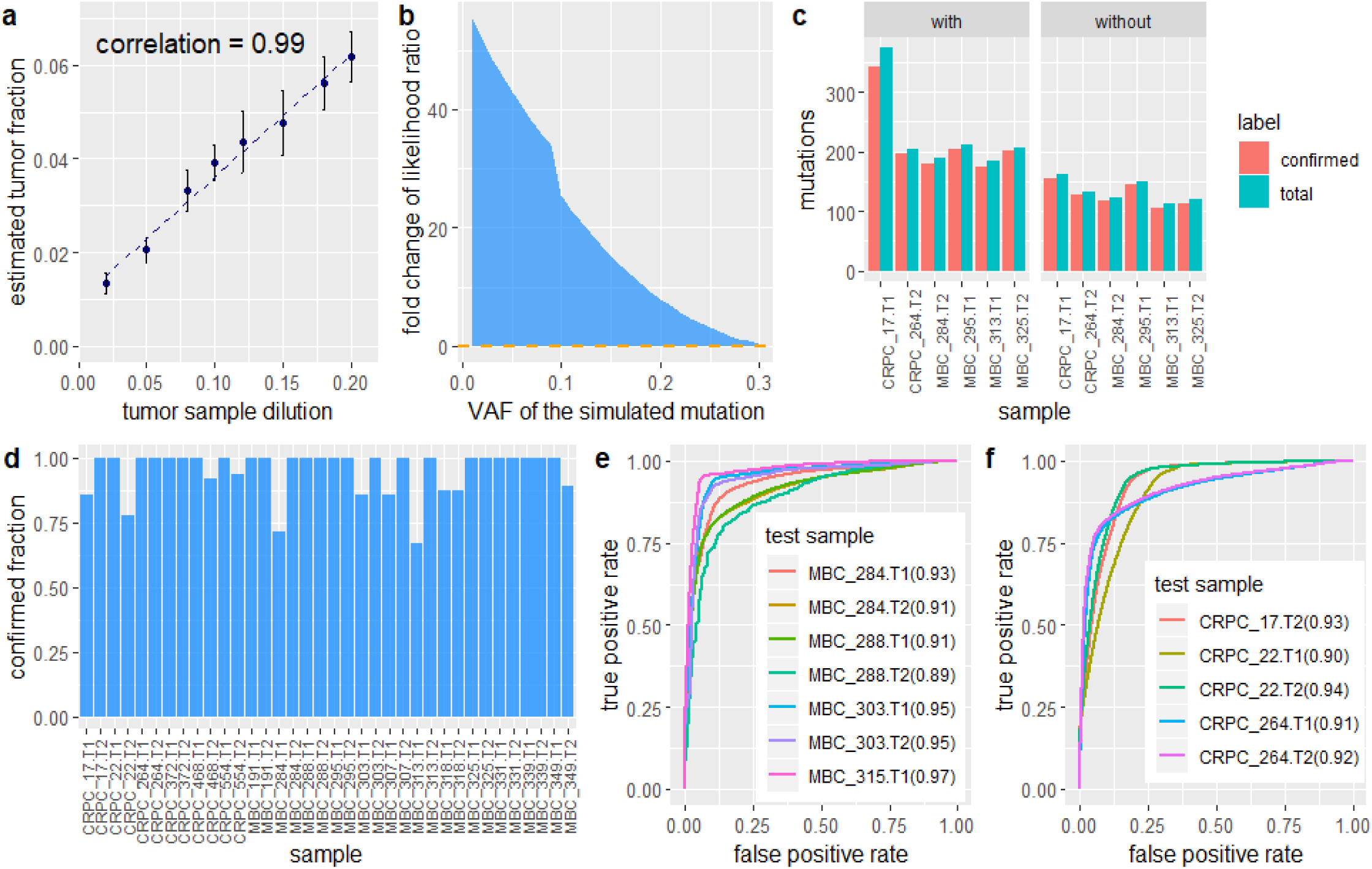
Experimental analysis of five new techniques. **a**, Performance of mutation cluster frequency estimation in terms of the correlation between the estimated tumor fraction and the true dilution ratio. This experiment uses simulated data based on WES of a single patient, with dilution ratios ranging from 2% to 20%. The points are the means ± s.d. of five independently generated datasets at each dilution. **b**, the fold change in the likelihood ratio between *cfSNV* models with and without a step to estimate the mutation cluster frequency, based on simulated mutations at different VAFs. **c**, Number of mutations detected with and without the iterative screening procedure. **d**, Confirmation rate of rescued mutations after adjusting conventional site-level post-filtration. **e-f**, Performance of read-level variant classifier on testing data. **e**. The ROC of applying the classifier to labeled data taken from seven cfDNA sequencing samples of four metastatic breast cancer patients. **f**. The ROC of applying the classifier to labeled data taken from five cfDNA sequencing samples of three metastatic prostate cancer patients. The numbers in parentheses indicate the area under curve (AUC) metric for each test sample.

**Fig. 5.**
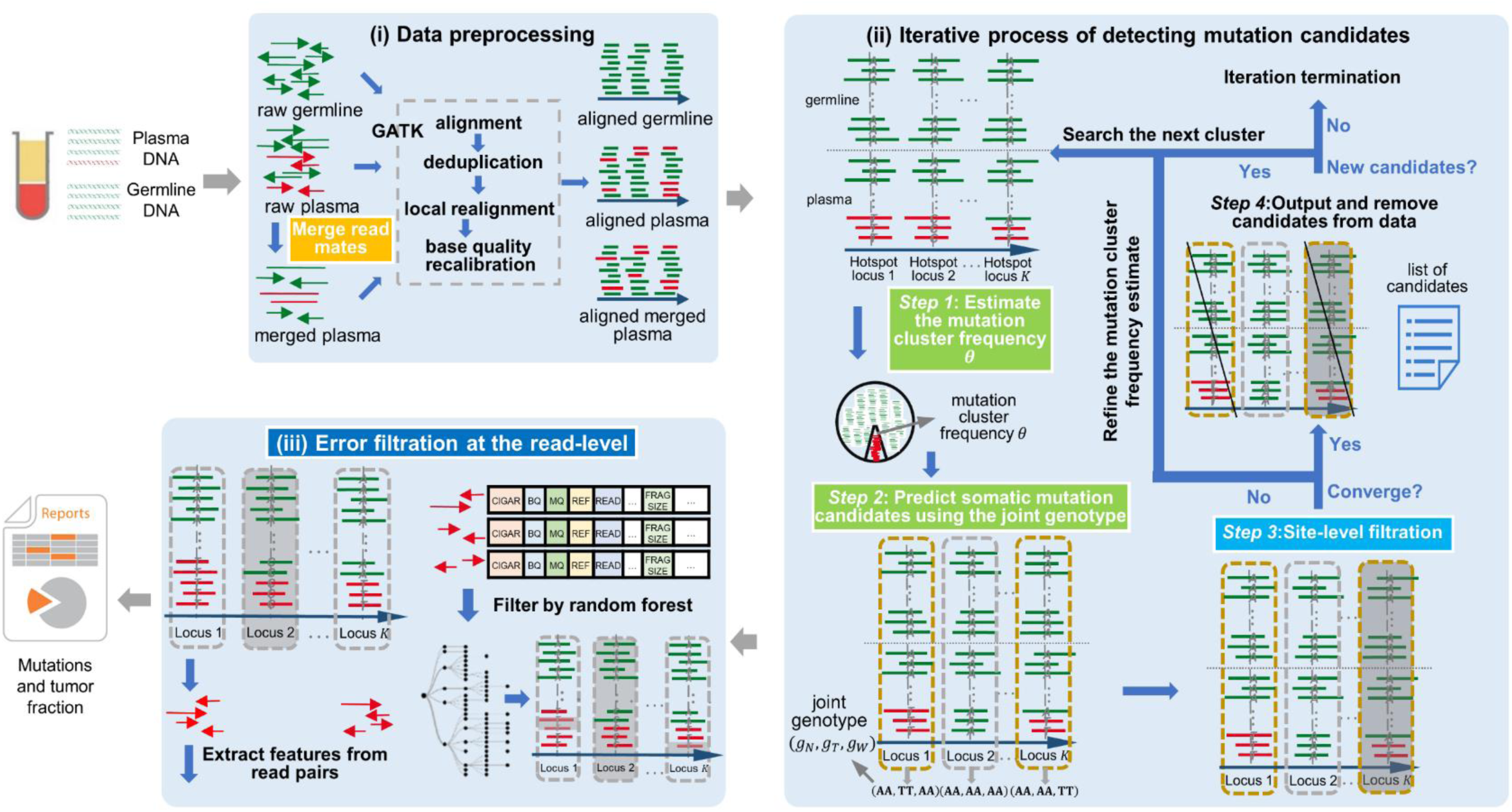
Workflow of *cfSNV*. *cfSNV* takes plasma DNA and germline DNA sequencing data as inputs. It first merges overlapping read mates in cfDNA sequencing data. The reads are processed using the GATK pipeline. After these steps, an iterative procedure estimates the mutation cluster frequency *θ* based on a set of carefully selected hotspots. Each iteration step determines the joint tumor-normal genotypes across sequencing regions, then eliminates somatic SNV candidates that fail essential filters based on site-level statistics (Methods). Mutation candidates are used as hotspot sites to refine *θ* and candidate detection until the frequency converges. SNV candidates from the previous iteration are output and masked before the next iteration. After all candidates are detected, a random forest classifier identifies raw read pairs with sequencing errors. Finally, somatic SNVs are reported only if enough variant supporting read pairs passed the random forest screening.

#### (iii) Enhance the sensitivity of mutation detection by an iterative process

We compared two versions of *cfSNV*, with and without the iterative process, on real data: four cfDNA samples whose prevalent mutation clusters have low prevalence (<20% estimated from *cfSNV* and *ichorCNA*). With the iterative process, *cfSNV* detected 1.41 to 1.73 times more confirmed mutations (true positives) than *cfSNV* without the iterative process (Fig. 4c). Both versions had high precision: namely, 95.3% and 95.0% for *cfSNV* with and without the iterative process respectively (Fig. 4c).

#### (iv) High confirmation rate of rescued mutations by cfDNA-specific post-filtration

Compared with the conventional post-filtration strategy, which models the randomness of variant-base positions on reads, our new filtration strategy rescues 1∼16 mutations (6.8 on average) per sample among the 36 plasma samples in this study. In 26 (69.4%) of the samples, 100% of the rescued mutations are confirmed in either the matched tumor biopsy or the other plasma sample (Fig. 4d).

#### (v) Machine learning approach to distinguish true mutations from sequencing errors in cfDNA reads

The out-of-sample data used to test the machine learning model are data from 7 MBC and 5 CRPC patients. We hand-labeled read pairs containing high-confidence mutations or sequencing errors, and applied the random forest classifier (Methods). Our method achieves an average AUC-ROC of 0.93 over the MBC cfDNA samples (Fig. 4e) and an average AUC-ROC of 0.92 over the CRPC cfDNA samples (Fig. 4f). This result implies that our machine learning model is non-specific to tumor types, and can easily be generalized to include samples from many kinds of tumors.

## Discussion

We presented a new computational framework, *cfSNV*, that sensitively detects low-frequency somatic SNVs in cfDNA sequencing data. *cfSNV* is equipped with a series of innovative techniques to address cfDNA-specific challenges (mixed tumor-/normal-derived cfDNA, low tumor-derived cfDNA fraction, high heterogeneity, and non-random fragmentation) and take advantage of cfDNA-specific features (high rate of overlapping reads, complete coverage of the mutation landscape). Specifically, (1) we designed a joint-genotype statistical model, parametrized by the mutation cluster frequency, to probabilistically deconvolute the mixture of tumor- and normal-derived reads in cfDNA data; (2) we developed an iterative approach to detect clusters of mutations with different variant allele frequencies; (3) we designed a data pre-processing step that exploits the overlapping read mates caused by short cfDNA fragments to improve data quality; (4) we developed a new procedure for filtering misalignment errors that accounts for the non-random fragmentation pattern of cfDNA; and (5) we developed a machine learning approach that incorporates the sequencing context to filter errors at the level of individual reads. These five techniques permit *cfSNV* to far outperform existing methods in sensitively and reliably detecting mutations in cfDNA, even with medium-coverage WES data.

The applicability of *cfSNV* to WES data enables cfDNA to be used in a wide variety of clinical applications. This paper presents one example of such an application: a novel and effective immunotherapy response measure that we name *truncal-bTMB*. This measure exploits cfDNA’s ability to provide comprehensive coverage of the mutation landscape and hence describe the complete clonal structure. The ability to obtain this information with a non-invasive blood test will greatly facilitate therapy prognosis and longitudinal monitoring.

## Methods

### Data collection

We collected WES data of 42 metastatic cancer patients from two sources: (1) The data for 41 patients were obtained from *Adalsteinsson et al*. [19] under dbGaP accession code phs001417.v1.p1. Each patient’s data include a WBC sample, a tumor biopsy sample, and one or two plasma cfDNA samples. Among the 41 patients, 18 have two plasma cfDNA samples. A patient (MBC_315) has her cfDNA sample sequenced with both WES and deep WGS. (2) The other patient was from *Butler et al*. [26], and her data were obtained under European Nucleotide Archive accession numbers ERS700858, ERS700859, ERS700860, and ERS700861. The data include a white blood cell sample, a primary breast cancer biopsy sample, a metastatic liver biopsy sample, and a plasma cfDNA sample.

### Human subjects

We collected blood samples, tumor biopsy samples and white blood cell samples from 30 non-small-cell lung cancer patients, who all provided informed consent for research use. The project was approved by the Institutional Review Boards (IRBs) of University of California, Los Angeles (IRB# 12-001891, IRB# 11-003066, and IRB# 13-00394).

### Plasma cfDNA whole exome sequencing (WES) library construction

For each of the 30 non-small-cell lung cancer patients, cfDNA was extracted from their plasma samples using the QIAamp circulating nucleic acid kit from QIAGEN (Germantown, MD). The cfDNA WES library was constructed with the SureSelect XT HS kit from Agilent Technologies (Santa Clara, CA) according to the manufacturer’s protocol. In brief, 10ng of cfDNA was used as input material. After end repair/dA-tailing of cfDNA, the adaptor was ligated. The ligation product was purified with Ampure XP beads (Beckman-Coulter, Atlanta, GA) and the adaptor-ligated library was amplified with index primer in 10-cycle PCR. The amplified library was purified again with Ampure XP beads, and the amount of amplified DNA was measured using the Qubit 1xdsDNA HS assay kit (ThermoFisher, Waltham, MA). 700-1000 ng of DNA sample was hybridized to the capture library and pulled down by streptavidin-coated beads. After washing the beads, the DNA library captured on the beads was re-amplified with 10-cycle PCR. The final libraries were purified by Ampure XP beads. The library concentration was measured by Qubit, and the quality was further examined with Agilent Bioanalyzer before the final step of 2×150bp paired-end sequencing by Genewiz (South Plainfield, NJ), at an average coverage of 200.

### The workflow of *cfSNV*

*cfSNV* takes the plasma DNA and germline DNA sequencing data of a patient as inputs, and detects SNVs using the three-step process described below (Fig. 5). The outputs at the end of the pipeline are the detected mutations and the tumor fraction.

#### (i) Data preprocessing

A short cfDNA fragment (one whose size is shorter than twice the sequencing read length) has overlapping read pairs, which introduce two important data preprocessing challenges: double-counting the overlapping regions and biasing variant allele frequencies. Discarding overlapping regions [13, 27, 28] not only wastes a large amount of sequencing data, but the opportunity to detect and suppress sequencing errors when two copies of the original DNA template are available. Therefore, in addition to the standard data preprocessing steps of alignment, deduplication, local realignment, and base quality recalibration, we perform an additional step: merging overlapping read mates. This new step occurs before the standard preprocessing pipeline (Fig. 5). It permits us to correct the read counts in overlapping regions, thereby removing the bias in variant allele frequencies from double-counting, and also detecting sequencing errors by comparing the context of the two cfDNA copies in the overlapping region. Later in the pipeline, inconsistent bases in the overlapping region are corrected to the base call with higher quality, while consistent bases are confirmed and assigned a high base quality. Specifically, we applied *FLASh* [29] for read mate merging. The parameters for *FLASh* were adjusted to accommodate the typical fragment lengths of cfDNA and read lengths in sequencing data. After running *FLASh* on the raw reads, those read mates that are likely to be overlapping are merged as single-end reads, while the rest of read pairs are treated as paired-end reads. We align paired-end reads and single-end reads separately to the hg19 human reference genome. We use *bwa mem* [30] to align the reads, and *samtools* [31] to sort them. Then we use *picard tools* [32] *MarkDuplicates* to remove duplicate reads resulting from PCR amplification. After this step, we add read group information to the bam file using *picard tools AddOrReplaceReadGroups*, and realign reads around indels using *GATK* [27, 28]. The target regions in realignment are identified through *GATK RealignerTargetCreator*, then reads around target regions are realigned using *GATK IndelRealigner*. Finally, base quality scores are recalibrated using *GATK BaseRecalibrator* and *PrintReads*.

#### (ii) Iterative process of detecting mutation candidates

As illustrated in Fig. 5, this process repeats a sequence of four steps until no more mutation candidates can be detected with confidence.

##### (Step 1) Estimating the mutation cluster frequency *θ* for the most prevalent mutation cluster

As the frequency of mutations in cfDNA are naturally clustered according to the clonal hierarchy [11, 18], we define a mutation cluster as a group of mutations with similar variant allele frequencies. The mutation cluster frequency *θ* is defined as the fraction of cfDNA carrying the mutations in the cluster, out of all cfDNA mapped to the same genomic positions. Due to the low amount of tumor-derived cfDNA in blood, individual sites may be covered by a very small number of tumor-derived cfDNA reads (or none), leading to highly uncertain estimates of the tumor-derived cfDNA fraction. Therefore, the first step is to identify sites across the genome that are highly likely to be mutated (called hotspots). Specifically, a locus is selected as a hotspot if it meets the following criteria: (a) both matched germline DNA and cfDNA sequencing data have adequate coverage (30 for germline, 80 for cfDNA in this study); (b) bases at the locus in matched germline DNA data contain only reference alleles; (c) the average sequencing error probability is less than the variant’s observed frequency; (d) reads in both matched germline DNA and cfDNA data have high mapping quality (≥ 20); (e) no strong strand bias is observed; and (f) enough variant supporting reads are observed in the cfDNA data (≥ 3). All hotspots are ranked by read coverage, variant allele frequency, and the counts of variant alleles in matched germline DNA data. Next, we estimate *θ* by maximizing the likelihood of observing the data at all hotspots P(X|*θ*), where X = (*X*_*1*_, *X*_*2*_, *⋯, X*_*r*_, *⋯*) is the cfDNA sequencing data and *X*_*r*_ represents all the information (such as sequence and base qualities) contained in a single read *r*. For each locus, we assume that reads are independently sampled from a cfDNA joint-genotype model denoted by the triplet *G* = (*g*_*T*_, *g*_*N*_, *g*_*W*_) where the subscripts *N, T* and *W* refer to normal-derived cfDNA, tumor-derived cfDNA and WBC DNA respectively. However, only the normal-derived cfDNA genotype *g*_*N*_ and tumor-derived cfDNA genotype *g*_*T*_ are utilized in this step, because the WBC genotype *g*_*W*_ is already controlled by hotspot selection (criterion b). All three genotypes are used in *(Step 2)* and *(Step 3)*, described below. Specifically, *g*_*W*_ becomes essential later in the process to remove germline mutations and WBC-derived somatic mutations (clonal hematopoiesis). Based on the independence of reads, the likelihood of a particular value for *θ* at a hotspot is calculated as the product of the probabilities of observing individual reads covering the hotspot, given the parameter *θ*. We express this relation as follows:

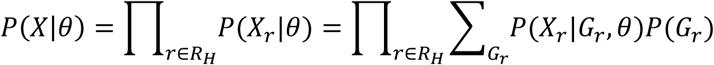

where *R*_*H*_ is the pool of reads covering a selected hotspot and *G*_*r*_ is the joint genotype at the hotspot covered by a read *r*. Note that sometimes a read *r* covers multiple hotspots, so *G*_*r*_ could be the combination of all hotspots covered by read *r*. Since an individual read is sequenced from either tumor-derived DNA (with probability *θ*) or normal-derived DNA (with probability *1* − *θ*), the likelihood of observing this read can be calculated using a probabilistic mixture model that describes the presence of two subpopulations:

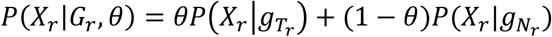

where 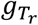 and 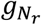 are the tumor-derived and normal-derived cfDNA genotypes of the hotspot on read *r*. The information contained in an aligned read *r* (*X*_*r*_) consists of base calls, base qualities and mapping qualities at hotspots in the read. So we can expand 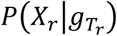 as follows:

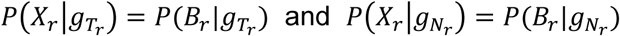

where *B*_*r*_ represents base calls at the hotspot on read *r*. The base quality and the mapping quality are embedded in the probability of sequencing error described below. The probability of error *ϵ* is calculated from the mapping quality *m* and the base quality *q*, as 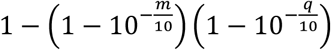 Assuming that all sequencing error directions have the same probability, the probability of observing a base call given genotype *g* can be calculated from the probability of error *ϵ*. So we have

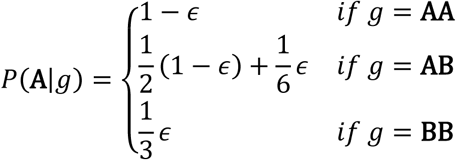

where **A** and **B** are the reference and non-reference alleles respectively. Based on the above formulation, an estimation of the mutation cluster frequency *θ* can be achieved by optimizing the likelihood *p*(*X*|*θ*) via the Expectation-Maximization (EM) algorithm or a simple grid search.

##### (Step 2) Predicting somatic mutation candidates using the joint genotype

After obtaining *θ*, we determine the variant status of a genomic position by finding the joint genotype that optimizes the posterior probability of reads at that position. As illustrated in Fig. 5(ii), for a given locus, we collect all reads aligned to the locus in both cfDNA data and the matched germline DNA data, then compute the posterior probability of each joint genotype from the observed reads. This probability can be modeled using a mixture model similar to that mentioned in *(Step 1)*. Subsequently, the joint genotype with the highest posterior probability is adopted as the prediction result at the locus. Somatic mutation candidates are then selected naturally following the inferred joint genotype. In this step, we use the matched germline data *X*_*W*_ from WBC and the cfDNA data *X*_*p*_ from plasma cfDNA, consisting of normal-derived cfDNA and tumor-derived cfDNA. For a specific locus, its joint genotype is determined as *G*_*MAp*_, the joint genotype that maximizes the posterior probability given the observed data and *θ*:

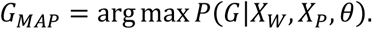

Using Bayes’ theorem, we have

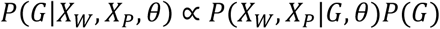

The probability of observing the data is the product of the probability of observing individual reads. So we have

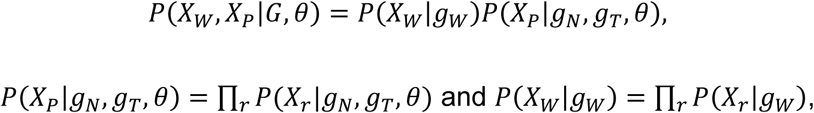

where *X*_*r*_ stands for a single read *r*. In the same way we calculate the likelihood of a given *θ*, we decompose *p*(*X*_*r*_|*g*_*N*_, *g*_*T*_, *θ*) and (*X*_*r*_|*g*_*W*_), and get

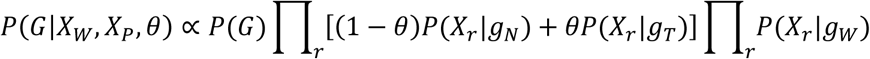

As the majority of normal-derived cfDNA comes from WBCs, we set the prior distribution of the joint genotype *G* as

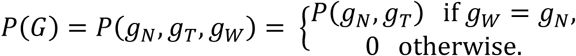

The joint distribution of the component (*g*_*N*_, *g*_*T*_) in joint genotype *G* has been defined in *JointSNVMix* [33]. It can also be calculated from public databases. Based on above formulation, the full joint genotype can be determined for every locus. By comparing the three components of the joint genotype with the highest posterior probability, we can determine whether the locus is a somatic mutation, a germline mutation, or a loss of heterozygosity (LOH) site. The somatic mutation loci are input as mutation candidates in the next filtration steps. The above model permits a probabilistic deconvolution of the normal and tumor signals in cfDNA. By incorporating the matched germline data (WBC) and the mutation cluster frequency *θ*, we separate the tumor-derived cfDNA from the total cfDNA at individual somatic SNV candidates, and thus enhance mutation detection (as shown in **Experimental analysis of five new techniques** (ii)).

##### (Step 3) Site-level filtration

To reduce false positives from mutation candidates, we investigate a set of site-level statistics in raw data and *FLASh*-processed data (i.e., both single-end reads from merged overlapping read pairs, and paired-end read pairs without overlapping regions). The site-level statistics used here include averaged base quality, averaged mapping quality, strand bias, depth of coverage, and nearby sequencing context (e.g. repeats and indels). Detailed descriptions and default thresholds for these site-level filters are listed in Supplementary Table 2. Based on the results from all filters, each mutation candidate is sorted into one of three categories: “pass”, “hold”, or “reject”. Candidates in the “pass” category pass all filters, so they are very likely to be mutations. Candidates in the “hold” category fail some non-essential filters, so we cannot determine whether they are mutations at this step. Candidates in the “reject” category fail at least one essential filter (e.g. averaged base quality), so they are regarded as false positives and removed from further analysis. The requirements for a variant to be classified as either “pass” or “hold”, are listed in Supplementary Table 2.

##### Iterating (Steps 1-3) to refine the mutation cluster frequency estimate

After *(Step 3)*, we select hotspots from the mutation candidates in the “pass” category to refine the *θ* estimation in *(Step 1)*. By repeating *(Steps 1-3)* for the same mutation cluster, we obtain a stable frequency estimate and a group of mutation candidates for this cluster. Convergence is reached when the difference between two consecutive *θ* estimations is less than 0.01. In our experiments with simulation data, convergence was usually reached after only two rounds (Supplementary Fig. 4). Thus, with just one iteration of *(Steps 1-3)*, we already accurately capture the most significant mutation cluster. In fact, our software offers both options: a quick version that performs only one round of estimation and candidate detection for each cluster, and a slow version that iterates until convergence for each mutation cluster.

##### (Step 4) Outputting and removing candidates from data

After obtaining somatic mutation candidates for the most prevalent mutation cluster, we output the mutation candidates in the “pass” and “hold” categories from *(Step 3)*. Then we remove the loci and data of these sites from the cfDNA data. After removal, we continue iterating from *(Step 1)* to identify the next most prevalent mutation cluster.

##### Termination criterion

Mutation clusters are detected one at a time, in decreasing order of their prevalence in cfDNA. The process ends when no mutation candidates are found following *(Step 4)* (i.e., the “pass” and “hold” categories are empty).

#### (iii) Error filtration at the read level

Site-level statistics provide some information on the difference between sequencing errors and true mutations, but are not adequate for error filtration in cfDNA. Due to the low tumor fraction and high heterogeneity of cfDNA, site-level frequency estimates are uncertain and unreliable for mutations with only a few supporting reads. To reduce the number of false positives among mutation candidates, we developed a machine learning filter to eliminate reads with sequencing errors at candidate sites, and remove SNV candidates whose count of “confirmed” supporting reads fails to pass a threshold (see details in Supplementary Table 3). Specifically, for each mutation candidate, we classify each of its supporting reads with a random forest model in order to distinguish sequencing errors from true variants. This model combines a variety of features (Supplementary Table 4) and automatically discovers statistical relationships among the features that reflect sequencing errors. It is worth noting that read pair statistics (e.g. fragment length and features of the read mate) are always among the most informative features of the random forest model. Since this error filtration method is applied at the read level, it vastly improves the precision of detecting low-frequency somatic mutations.

Although this read-level filter can be performed at any step of the method (e.g., after alignment or during the iterations), we prefer to perform it at the end of the *cfSNV* workflow in order to save computing time and resources. Generally, the later this step is performed, the fewer sequencing reads need to be inspected for errors: based on our observations of the real data, for each read that needs to be inspected at the end of the process, at least 50 reads would need to be inspected at the beginning.

To train the random forest model, we used four WES sequencing datasets from the same cancer patient (MBC_315): two cfDNA sequencing datasets, a WBC sequencing dataset, and a tumor biopsy sequencing dataset. As the two cfDNA sequencing datasets are on the same cfDNA sample, we can treat them as technical replicates and label their read pairs by their concordance. The training data are the supporting cfDNA read pairs at known mutation/error sites, and labeled as containing mutations or errors. Mutation sites are defined as the collection of common germline mutations detected using *Strelka2 germline* [35] from all four datasets. In addition, common somatic mutations were detected using *Strelka2 somatic* and *MuTect* [13] from two cfDNA-WBC pairs (cfDNA data vs. WBC data) and one tumor-WBC pair (tumor data vs. WBC data). Error sites are defined as sufficiently covered sites (> 80x) with only one high-quality non-reference read (base quality ≥ 20 and mapping quality ≥ 40) in all four datasets. All labeled read pairs were extracted from raw cfDNA data using Picard *FilterSamReads* (Supplementary Table 4). Different features were constructed from the overlapping read pairs and the non-overlapping read pairs (Supplementary Table 5). All categorical features were expanded using one-hot encoding. We set the parameters of the random forest model as follows: (1) the number of decision trees is 100, (2) the maximum tree depth is 10, (3) imbalanced classes were handled by setting the class weights with option “balanced”, and (4) other parameters were left at their default values. Two random forest classifiers (for overlapping read pairs and non-overlapping read pairs) were trained on read pairs extracted from the WES data (SRR6708941) using RandomForestClassifier from the python library scikit-learn [36]. Read pairs from SRR6708920 were only used for validating the model. The trained classifiers are saved in the *cfSNV* code package (upon the request).

### Truncal-bTMB measure

Somatic SNVs are annotated using *snpEff*. Nonsynonymous mutations and high-impact mutations are treated the same in *snpEff* results. As the VAF in cfDNA reflects the clonality of a mutation, we treat mutations as truncal mutations if their VAF is greater than a threshold; otherwise the mutation is a branch mutation. The threshold is defined as 60% of the average VAF of the 5 most frequent mutations. The truncal-bTMB measure can then be calculated as the sum of the observed VAFs for all truncal nonsynonymous mutations.

### Additional validation data for random forest classifier

To further test the random forest classifiers, we generated data from other patients with metastatic breast or prostate cancer (Supplementary Table 1). For each patient, we obtained WES data of a WBC sample, a tumor biopsy sample, and plasma samples from two different time points. To generate the testing data and label the individual reads, we used the same procedure as described in **Error filtration at the read level** for producing the training data.

### Simulation with *BAMSurgeon* to evaluate precision and sensitivity

To evaluate the performance of *cfSNV*, we employed *BAMSurgeon* [37] to generate simulation data by inserting individual mutations at different allele frequencies. The input to *BAMSurgeon* was a pool of germline DNA data from eleven breast cancer patients (MBC_331, MBC_335, MBC_299, CRPC_22, MBC_295, CRPC_264, CRPC_342, CRPC_17, MBC_339, MBC_313, and MBC_321) [19]. The mean target coverage of the pooled sample reached 2000x. The program attempted to insert 900 somatic SNVs with different variant allele frequencies: 100 at 5%, 100 at 3%, 100 at 1%, 100 at 0.8%,100 at 0.5%, 200 at 0.3%, and 200 at 0.1%. A total of 561 mutations were successfully inserted. The other 339 mutations failed to insert into the sequencing data because their assigned VAF was incompatible with the sequencing depth in the original data, e.g. 1% VAF among 10 reads. We evaluated the performance of *cfSNV, MuTect* (disabling the contamination filter and testing different levels of the tumor_lod parameter) and *Strelka2* (default parameters, with enabled and disabled filters) on this simulation dataset by comparing the ground truth to the final variant reports. *MuTect* performed best when “tumor_lod” was set to 5, so we only report its results for this setting.

### Mutation concordance between tumor biopsy and plasma samples

To validate our method on real data, we examined mutation concordance between a tumor biopsy sample and the plasma samples. This analysis involves twelve patients with metastatic breast cancer and six patients with metastatic prostate cancer [19]. Each patient had a tumor biopsy sample, a WBC sample, and plasma samples from two different time points, all processed with WES. Mutations called from one plasma sample were checked in the raw sequencing data of the matched tumor biopsy sample and the other plasma sample. A somatic SNV is confirmed if there are at least three reads supporting the variant allele in the matched tumor biopsy sample or at least three reads supporting in the other plasma sample. A somatic SNV is not confirmed when the mutation has power at least 0.9 and fewer than 3 alternative reads [19].

### Comparison with *MuTect* and *Strelka2* on real cfDNA data

We compared our method to two state-of-the-art methods, *Mutect* ad *Strelka2*. The same validation analysis was conducted for both methods on the same samples. Both tools were run with their default parameters unless otherwise noted in the text. The same confirmation process described in **Mutation concordance between tumor biopsy and plasma samples** was conducted for somatic SNVs detected by *MuTect* and *Strelka2*.

### Calculation of TMB and bTMB

For tissue biopsy samples, we called their somatic SNVs using *VarScan2* [38] with default parameters. Mutations were filtered if the depth of coverage was lower than 35, if the count of variant supporting reads was lower than 25, or if the observed variant allele frequency was lower than 0.05. The remaining mutations were annotated using *snpEff* [39]. TMB was calculated as the number of nonsynonymous SNVs. For plasma samples, we called somatic mutations using *cfSNV* and annotated them using *snpEff*. We calculated traditional bTMB as the count of all nonsynonymous mutations with VAF ≥ 0.15.

### Simulation with BAMSurgeon to evaluate the accuracy of intelligent search

We used *BAMSurgeon* to generate simulation data. The input to *BAMSurgeon* was the WBC sequencing data from MBC_299. The program attempted to insert 300 mutations at three different VAF levels: 50 mutations at 20%, 150 mutations at 8%, and 100 mutations at 2%. Five simulated samples with the same settings were generated.

### Generating spike-in simulation data to validate the mutation cluster frequency estimates

To evaluate the accuracy of our mutation cluster frequency estimation, we generated spike-in simulation data by mixing the primary tumor sequencing data (ERS700859) and the WBC sequencing data (ERS700858) of a metastatic breast cancer patient, at varying concentrations of cfDNA reads (from 2% to 20% in eight steps). Five independent mixtures are generated at every concentration. Each spike-in sample contains a total number of randomly sampled reads equivalent to 170x coverage of the targeted regions. The coverage of the targeted regions is limited by the number of sequencing reads in the original data.

### Impact of the mutation cluster frequency on the model-to-data fitness at a single simulated mutation

The model-to-data fitness is evaluated using the likelihood ratio *L*_*θ*_, the ratio between the maximum likelihood of a somatic-mutation joint genotype (i.e., homozygous and heterozygous genotypes) and the maximum likelihood of a non-somatic-mutation joint genotype (other joint genotypes) given an *θ*. Since we screen mutation candidates based on the joint genotype estimated at each position, this likelihood ratio reflects the ability of *cfSNV* to detect a somatic mutation candidate. We explore the theoretical properties of this likelihood ratio using simulated mutations, which consist of randomly generated base quality values, mapping quality values and a corresponding list of base calls reflecting the VAF. To compare the fitness of the model with and without *θ*, we calculated the value of *L*_*θ*_*/L*_1_.

### Impact of the mutation cluster frequency on real patient data

To test the impact of estimated mutation cluster frequency on real patient data, we selected four samples whose prevalent mutation clusters have low prevalence <20% estimated from *cfSNV* and *ichorCNA*. We ran *cfSNV* on the four samples using both a predetermined value of *θ* (0.2, 0.5, 0.8, and 1.0) and the estimated *θ* of the most prevalent mutation cluster in the sample. When we set *θ* to 1.0, the candidate screening model is the same as the regular joint genotype model for solid tumor samples, which is equivalent to a model that does not incorporate the estimated mutation cluster frequency. In this simulation, we also disabled the iterative procedure to converge on the best value of *θ*, so the candidate screening only took place at the given *θ*.

### Rescuing mutations from conventional post-filtration

We remove the conventional clustered read position filter entirely. Instead, to remove misalignment artifacts, we implemented a new filter that simultaneously checks the co-occurrence of candidates on the reads. If several candidates exclusively co-occur on the variant supporting reads, we regard them as artifacts from misalignment (Supplementary Table 2). A mutation is called “rescued” if it is reported by *cfSNV* but would be filtered by conventional methods due to clustered read position. For each rescued mutation, the same confirmation process described in **Mutation concordance between tumor biopsy and plasma samples** was conducted. The fraction of confirmed rescued mutations among all rescued mutations was calculated for every sample. Indeed, we were able to confirm that for some rescued mutations, the variant bases are more clustered in cfDNA reads than in solid tumor samples (Supplementary Fig. 5), validating our rationale.

## Data Availability

Raw data related to this manuscript can be provided upon request. The links to public data are included in Methods.

